# Congruence of Health-related Quality of Life Assessment between Patient-Physician and Patient-Caregiver Dyads

**DOI:** 10.1101/2020.09.29.20204404

**Authors:** Chia-Chun Tang, Hsi Chen, Wei-Wen Wu, Jaw-Shiun Tsai

**Author notes:** Corresponding Author: Chia-Chun Tang, PhD, School of Nursing, College of Medicine, National Taiwan University, 10051 No. 1, Sec. 1, Jen-Ai Road, Taipei, Taiwan. Author Contributions: Conceptualization, C.T.; methodology, C.T.; software, C.T. and H.C.; validation, C.T., H.C., W.W., J.T.; formal analysis, H.C. and C.T.; investigation, H.C.; resources, J.T.; data curation, C.H.; writing—original draft preparation, C.T. and H.C.; writing—review and editing, C.T., H.C.,W.W., and J.T.; visualization, W.W.; supervision, W.W. and J.T.; project administration, C.T.; funding acquisition, C.T. All authors have read and agreed to the published version of the manuscript.

## Abstract

**Background:** Exploring the congruence of health-related quality of life (HRQOL) evaluation provides valuable information regarding whether proxies can accurately reflect patient perceptions and the quality of symptom communication. It is particular important in advanced cancer population who experience poor HRQOL and have deteriorated ability to express their feeling. The main purpose of this study was to investigate the congruence of HRQOL reports between patient-physician and patient-caregiver dyads and to determine the association of variables, if any, with the congruence between dyads.

**Methods:** This observational study with a cross-sectional design first approached physicians who provided care for patients with advanced cancer at the participating institution. Then, participated physicians’ patients and their caregivers were recruited. All participants were required to independently fill out an HRQOL questionnaire during their outpatient visits. Descriptive statistics, weighted kappa, Wilcoxon signed-rank test, and linear regression were employed for data analysis.

**Results:** A total of 52 patient-physician and 27 patient-caregiver dyads were examined. Patients suffered from considerable problems in all three HRQOL domains: symptom, functioning, and overall HRQOL. The patients’ level of agreement was moderate with the caregivers and fair with the physicians. A significant relationship was found between several patient-related variables and disagreement.

**Conclusion:** The study’s patient group experienced a compromised HRQOL, warranting immediate attention. When there are barriers to obtaining a patient’s self-repot, clinicians may consider caregivers as reasonable source. Some patients with special characteristics may need additional attention because their problems may be at greater risk of being overlooked.

## Background

An individual is the most ideal and reliable source of their health-related quality of life (HRQOL). However, it is common to determine the status of terminally ill patients based only one the evaluation of their proxies owing to the patients’ deteriorating physical and psychological conditions. Hence, examining the extent to which these proxy raters can be relied on and the factors that may affect their evaluation is imperative. In addition, exploring the degree of congruence between patient’s and healthcare providers’ evaluation provides clues to the communication quality.

Studies focusing on patient-caregiver agreement of the evaluation regarding various domains of HRQOL have increase significantly over the past two decades. Experts have investigated the agreement of HRQOL evaluation in diverse population, such as elderly, patients with cancer or heart failure, and hospice patients ^1-5^. Some studies addressed several domains of HRQOL together, while others have only focused on single symptom, such as pain. The results have been mixed - several studies have suggested that caregivers’ report of cancer patients’ HRQOL is close to patients’ self-report ^1 2 6-8^; however, other studies have reported different evaluations between the caregivers and the patients ^3 9^. A systematical review has recommend that caregivers estimate the observable symptoms accurately but not the subjective experiences ^10^. Although the agreement levels have been varied, many studies agree that caregivers generally overestimate the patients’ problems ^2 3 8 11^.

Researchers have found that healthcare providers have different tendencies as compared to caregivers; healthcare providers’ perspectives deviated more from the patients’ ^12-14^, and often underestimated their problems ^15-19^. Patient factors such as gender, age, marital status, history of drug abuse, first-time admission, severity of functional impairment or symptoms, and anxiety are all possible influences that affect the agreement between proxies ^11 13 14 16 19 20^. As the evidence regarding the level of congruence between patients and proxies has been developing, more questions have emerged. First, although symptoms, by definition, are “perceived indicators of change in normal functioning as experiencing by patients ^21^,” symptom report and evaluation is essentially a communication process shared among patients, caregivers, and healthcare providers. Thus, the discrepancy between evaluations is not only associated with how well proxies can capture patients’ feelings, but also indicates the quality of their communication. However, most relevant studies have not addressed any issues regarding this communication. Study designs that specify the timings between symptom evaluation and the related communication can provide hints regarding the quality of communication based on symptom agreement. For example, if the low congruence level is detected immediately after symptom discussion, it may suggest existing problems related to communication. Second, as current evidence was primarily based on data obtained from European and North America populations, it is important to examine the phenomenon in other populations and healthcare systems.

Accordingly, the current study was constructed in outpatient department (OPD) settings in Taiwan to (1) examine advanced cancer patients’ various domains of HRQOL as reported by patients, caregivers, and physicians, (2) to investigate the congruence level of HRQOL reports between patient-physician dyads and patient-caregiver dyads, and (3) to determine if any patient variables are associated with the congruence level between dyads. Collecting data in OPD settings allowed us to emphasize the timings between evaluation and communication which was described in the following method section.

## Method

### Study Design

This was an observational study with a cross-sectional design. Physicians taking care of patients with solid tumor at a medical center in Northern Taiwan were first recruited through personal contacts. Once the physician agreed to participate, a research assistant consulted a participating physician to identify potentially eligible patients. The inclusion criteria for the potentially eligible patients included consulting a participating physician, being diagnosed with advanced solid cancer (TNM stage III or IV), aged 20 years or older, and being able to communicate in Chinese or Taiwanese were approached. However, patients who did not experience any symptoms or were hospitalized at recruitment were excluded. Simultaneously, adult caregivers who accompanied the participating patients to their OPD visits were also invited to participate in the study. Both participating patients and caregivers had to independently fill in their own questionnaires either shortly before or after their OPD visits. On the other hand, physicians were required to complete the same questionnaire for each participating patient immediately after their OPD discussion. The data collection period was from December 2018 to December 2019. This study was approved by the National Taiwan University Hospital Human Subjects Office Institutional Review Board.

### Instrument

The European Organization for Research and Treatment of Cancer quality of life questionnaire (EORCT-QLQ C30, Taiwan Chinese version) is a 30-item questionnaire evaluating cancer patients’ HRQOL according to three domains: symptom, functional impairment, and the overall HRQOL. While items in the overall HRQOL domain are rated on a 7-point Likert scale, other items are rated on a 4-point Likert scale. The final scores for each domain are standardized and transformed to a range of 0-100. Higher scores indicate better conditions in HRQOL domain, but worse problems in symptom and functioning domains. The EORTC QLQ-C30 has been verified and widely applied in health-related research; the Taiwan Chinese version has been tested and shows good reliability and validity ^22 23^. The reported Cronbach’s alpha coefficient for most items has been above 0.70 across studies. In this study, the Cronbach’s alpha for the responses of patients, caregivers, and physicians was 0.84, 0.92, and 0.93, respectively.

### Statistical Methods

Descriptive statistics were used to analyze the demographic data and score for each questionnaire item. Then, the score of each item was further compared to a clinically significant value to calculate the level of symptom burden ^24^. The patient’s level of symptom burden was the number of items that they rated as equal to or greater than the clinically significant value.

The congruence between dyads were analyzed from two aspects: individual and group. The individual aspect emphasized the congruence of each dyad, while the group aspect focused on the overall congruence between all patients and proxies and whether these differences reached statistical significance. Specifically, weighted kappa was employed to assess the data from the individual aspect what categorized congruence into six levels ^25^: no agreement (*k*=0), none to slight (*k*=0.01-0.20), fair (*k*=0.21-0.40), moderate (*k*=0.41-0.60), substantial (*k*=0.61-0.80), and almost perfect (*k*=0.81-1.00). A sample size of 32 for patient-physician dyads and 20 for patient-caregiver dyads were determined based on Bujang et. al.’s guideline of the kappa agreement test ^26^. We further calculate sum score differences of each dyad (physician or caregiver score minus patient score) to reflect the proportion of complete agreement (score=0), overestimation (score >0), or underestimation (score<0). With regard to the group aspect, the Wilcoxon signed-rank test was applied ^27^.

To explore whether there was any variable associated with incongruence in HRQOL evaluation, linear regression was performed. The dependent variables, incongruence in each domain, were the total sum score difference of each domain. The independent variables that could potentially affect congruence were identified through a literature review. Software such as Microsoft Excel (2016), IBM SPSS 21, RStudio, and Octave were used for statistical analysis and to generate figures.

## Results

A total of 52 patient–physician and 27 patient–caregiver dyads were included in the analysis. Among the eight physicians approached, six agreed to participate, including five males and one female (mean age=42.67; SD=5.5). Their average years of practice was 13.5 (SD= 3.77), either as medical oncologists (n=3) or hospice specialists (n=3). One hundred and twenty-four patients consulting the six participating physicians were approached, and 66 patients (53%) agreed to participate. There was no significant difference in age and sex between those who agreed and refused to participate; however, patients diagnosed with stage IV cancer were more likely to participate as compared to patients with stage III diagnosis *(p* = .014). Finally, 52 patients (78.78%) with 10 different types of cancer diagnoses completed the study (mean age = 61.62, SD = 12.12). The majority were male (80.8%), married (73.1%), and unemployed (55.7%). Thirty-five (67%) were recruited from the oncology OPD and 17 (32.6%) from the hospice OPD. The reasons for not completing the study were: deteriorated physical or psychological condition, loss of OPD follow-up or not planning to follow up at OPD, and lack of symptoms. Twenty-seven caregivers of the participating patients, including 24 females and 3 males, agreed to participate in the study. Their relationships with the patients were of partners (n = 16, 59.3%), children (n = 8, 29.6%), parents (n = 2, 7.4%), and siblings (n = 1, 3.7%). Their mean age was 55.22 years (SD = 9.28).

### Descriptive Data of HRQOL

Based on the patients’ perspectives, the top four symptoms that received the highest median scores were fatigue, appetite loss, insomnia, and financial difficulties with medians of 44.44, 33.33, 33.33, and 33.33, respectively. The median burden level was five, indicating that the majority of patients had at least five symptoms that reached clinical importance. The median scores for the social, physical, role, cognitive, and emotional functions were 66.66, 73.33, 83.33, 83.33, and 83.33, respectively. The median score for the overall HRQOL was 50.

On the other hand, both physician and caregiver ratings for HRQOL (median = 41.66 for both groups) were lower than the patients’ ratings. Physicians’ and caregivers’ evaluation of functions were similar to the patients’ self-ratings, except for role function and emotional function that had medians of 66.66 and 75, respectively, for both physician and caregiver groups. With regard to the symptoms, the physicians believed that pain, insomnia, fatigue, appetite loss, and constipation were all considerable problems for patients (median = 33.33 for all five symptoms). While most of the caregivers’ evaluations of symptoms were similar to that of the physicians, their ratings showed that insomnia (median = 66.66) and fatigue (median = 44.44) were the two most severe symptoms. Similar to the patients, the caregivers also highlighted financial difficulties (median = 33.33); however, the physicians considered it to be a minor problem (median = 0). The quartiles of scores for the three domains are listed in Table 1.

**Table 1.**
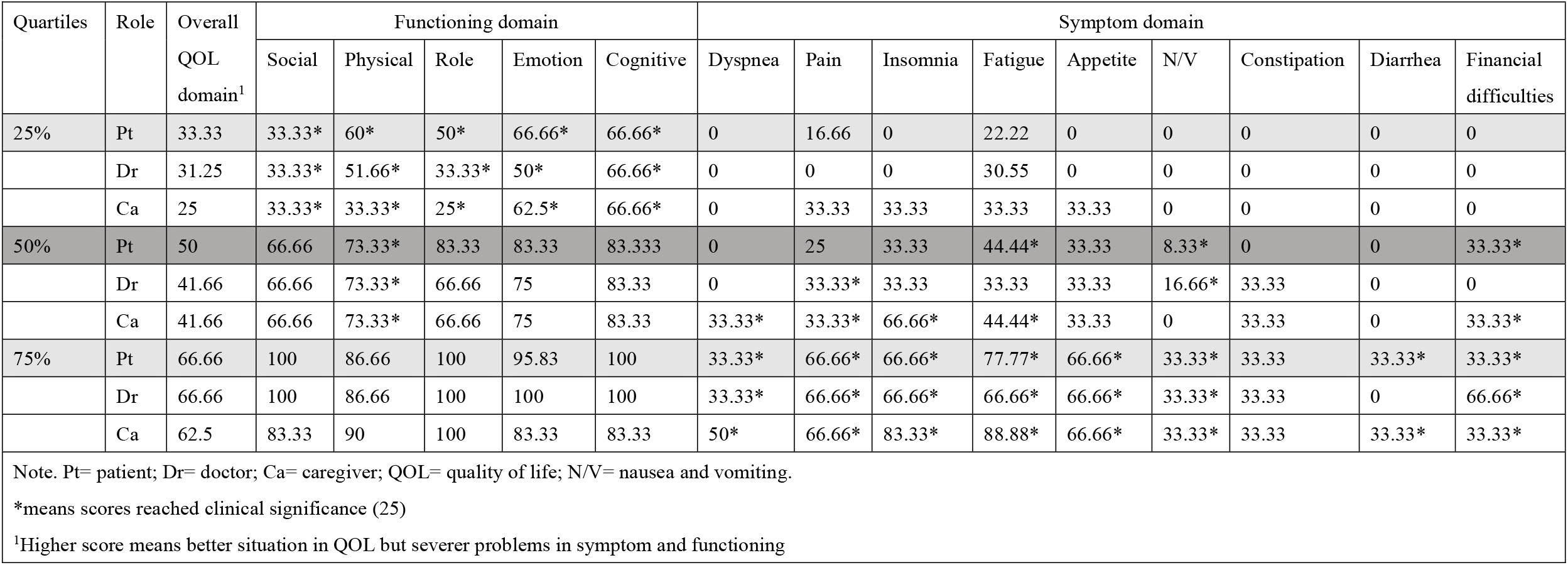
Prevalence and intensity of symptoms, functioning status, and quality of life as rated by patients, physicians, and caregivers

### Congruence of the Assessment of HRQOL between Dyads

The following paragraphs present the congruence of evaluation according to different dyads (i.e., patient–physician and patient–caregiver dyads) and aspects (i.e., individual and group).

#### Patient–physician dyads

From the individual aspect, patients and physicians had a fair level of agreement on majority of the items such as problems in role, emotional, cognitive, and social functioning, nausea and vomiting, insomnia, appetite loss, constipation, and diarrhea. However, their HRQOL ratings for problems in physical functioning, fatigue, pain, and dyspnea reached moderate agreement. Patient and physician dyads had none to slight agreement on financial difficulties (Figure 1). Physicians underestimated the patients’ HRQOL, problems in social functioning, fatigue, insomnia, and pain. However, they overestimated the problems in role, physical, and emotional functioning (Table 2). When assessing the congruence of symptom evaluation from the group aspect, patients and physicians had significantly different ratings for diarrhea and financial difficulties (Figure 1)

**Table 2.** Under- and Overestimation of Symptoms, Functioning, and Health Related Quality of Life by Physicians and Caregivers as Compared to Patients

| Physician ratings compared to patient ratings (n=45-52 pairs<sup>1</sup>) |  |  |  |  |  |  |  |  |  |  |  | Symptoms, Functioning<br>Impairment, and Health Related<br>Quality of Life | Caregiver ratings compared to patient ratings (n=25-27 pairs<sup>1</sup>) |  |  |  |  |  |  |  |  |  |  |  |
| --- | --- | --- | --- | --- | --- | --- | --- | --- | --- | --- | --- | --- | --- | --- | --- | --- | --- | --- | --- | --- | --- | --- | --- | --- |
| Physician Underestimation |  |  |  |  | Complete<br>Agreement |  | Physician Overestimation |  |  |  |  |  | Caregiver Underestimation |  |  |  |  | Complete<br>Agreement |  | Caregiver Overestimation<sup>2</sup> |  |  |  |  |
| -4 | -3 | -2 | -1 | % | 0 | % | +1 | +2 | +3 | +4 | % |  | -4 | -3 | -2 | -1 | % | 0 | % | +1 | +2 | +3 | +4 | % |
| 6 | 5 | 4 | 9 | 46.2 | 8 | 15.4 | 4 | 11 | 2 | 3 | 39.5 | Health Related Quality of Life | 2 | 2 | 5 | 3 | 44.4 | 7 | 25.9 | 4 | 3 | 1 | 0 | 29.6 |
| 4 | 2 | 11 | 5 | 45.8 | 11 | 22.9 | 5 | 4 | 2 | 4 | 31.3 | Social Functioning | 1 | 1 | 5 | 4 | 40.7 | 7 | 25.9 | 2 | 3 | 1 | 3 | 33.3 |
| 6 | 5 | 5 | 7 | 44.2 | 8 | 15.4 | 9 | 7 | 1 | 4 | 40.4 | Fatigue | 0 | 0 | 3 | 7 | 37 | 4 | 14.8 | 7 | 2 | 2 | 2 | 48.1 |
| 0 | 1 | 1 | 18 | 40.8 | 11 | 22.4 | 14 | 3 | 1 | 0 | 36.7 | Insomnia | 0 | 0 | 0 | 6 | 22.2 | 9 | 33.3 | 9 | 2 | 1 | 0 | 44.4 |
| 1 | 2 | 6 | 11 | 38.5 | 17 | 32.7 | 10 | 1 | 3 | 1 | 28.8 | Pain | 0 | 1 | 0 | 3 | 14.8 | 7 | 25.9 | 8 | 5 | 1 | 2 | 59.2 |
| 0 | 1 | 5 | 12 | 34.6 | 18 | 34.6 | 12 | 3 | 1 | 0 | 30.8 | Appetite Loss | 0 | 0 | 1 | 7 | 29.6 | 14 | 51.9 | 1 | 3 | 1 | 0 | 18.5 |
| 0 | 0 | 1 | 11 | 23.1 | 37 | 71.1 | 3 | 0 | 0 | 0 | 5.8 | Diarrhea | 0 | 0 | 0 | 5 | 18.5 | 16 | 59.2 | 6 | 0 | 0 | 0 | 22.2 |
| 0 | 0 | 3 | 6 | 17.3 | 29 | 55.8 | 7 | 7 | 0 | 0 | 26.9 | Constipation | 0 | 0 | 0 | 5 | 18.5 | 19 | 70.4 | 3 | 0 | 0 | 0 | 11.1 |
| 0 | 1 | 4 | 11 | 33.3 | 25 | 52 | 6 | 1 | 0 | 0 | 14.6 | Financial Difficulties | 0 | 0 | 2 | 2 | 14.8 | 14 | 51.9 | 8 | 1 | 0 | 0 | 33.3 |
| 0 | 0 | 2 | 45 | 26.9 | 24 | 46.1 | 10 | 3 | 1 | 0 | 26.9 | Dyspnea | 0 | 0 | 1 | 2 | 11.1 | 15 | 55.6 | 5 | 3 | 1 | 0 | 33.3 |
| 1 | 2 | 3 | 10 | 34 | 21 | 44.7 | 7 | 3 | 0 | 0 | 21.3 | Cognitive Functioning | 0 | 0 | 2 | 2 | 14.8 | 13 | 48.1 | 6 | 1 | 3 | 0 | 37 |
| 1 | 0 | 4 | 8 | 25 | 20 | 38.5 | 7 | 7 | 2 | 3 | 36.5 | Nausea and Vomiting | 0 | 0 | 4 | 3 | 25.9 | 12 | 44.4 | 5 | 1 | 1 | 1 | 29.6 |
| 0 | 3 | 8 | 3 | 28 | 12 | 24 | 5 | 11 | 2 | 6 | 48 | Role Functioning | 0 | 0 | 2 | 3 | 19.2 | 10 | 38.5 | 3 | 5 | 0 | 3 | 42.3 |
| 4 | 3 | 5 | 6 | 40 | 8 | 17.8 | 6 | 1 | 4 | 8 | 42.2 | Physical Functioning | 2 | 0 | 1 | 2 | 20 | 5 | 20 | 6 | 5 | 0 | 4 | 60 |
| 4 | 3 | 7 | 4 | 37.5 | 10 | 20.8 | 7 | 2 | 3 | 8 | 41.7 | Emotional Functioning | 1 | 1 | 2 | 2 | 23.1 | 5 | 19.2 | 3 | 5 | 3 | 4 | 57.7 |

*Note.* The comparison of the dyad's rating was counted by sum score differences (caregiver score minus patient score) on functional impairment/ symptoms/ Health Related Quality of Life. The grey area indicates where the majority of proxies underestimate, overestimate, or agree with the patients
<sup>1</sup> The pairs were eliminated if either patient, physician, or caregiver in that pair has an incomplete data. The original eligible pairs were 52 pairs for patient-physician dyads and 27 pairs for patient-caregiver dyads.
<sup>2</sup> Overestimated for all items suggest that the proxy rated the problem severer than the patient, except for Health Related Quality of Life ratings (overestimated for Health Related Quality of Life means that proxy perceived a better condition than patient's perception)

**Figure 1.**
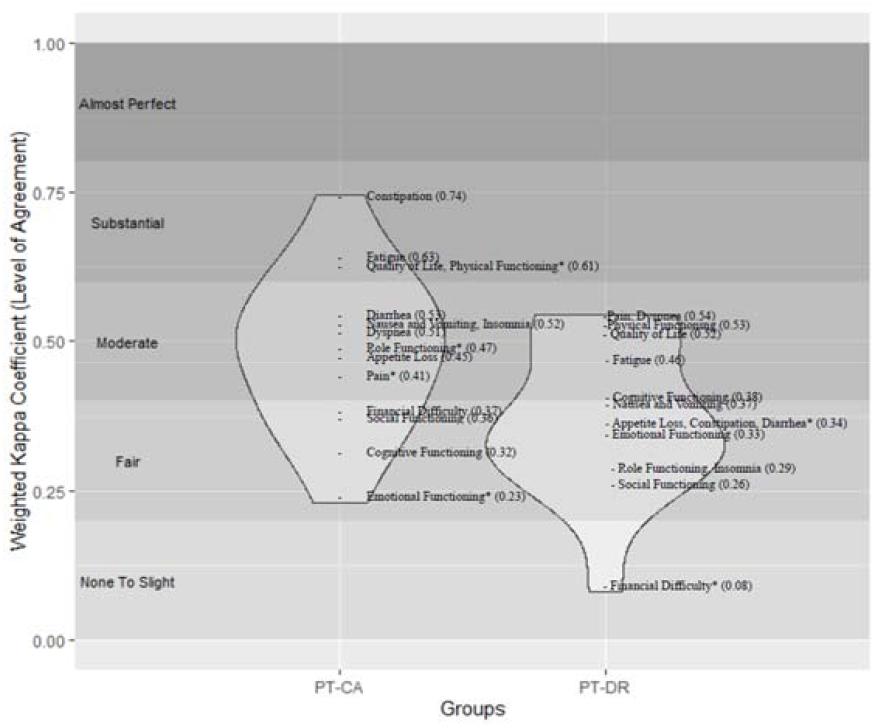
Level of Agreement for Patient-Caregiver Dyads and Patient-Physician Dyads: Numbers in parentheses represent weighted kappa coefficient between patients and caregivers and patients and physicians, the “*” sign implies significant difference on rating between the two (Wilcoxon signed-rank test). The black and white gradient background shows the interpretation of weighted kappa coefficient (level of agreement) as suggested by Landis and Koch’s (1977).

#### Patient–caregiver dyads

From the individual aspect, patients and caregivers had a moderate level of agreement on the majority of items such as problems in role functioning, nausea and vomiting, pain, dyspnea, insomnia, appetite loss, and diarrhea (Figure 1). Moreover, their ratings of HRQOL, physical functioning, fatigue, and constipation reached a substantial level of concordance. On the other hand, they had a fair level of agreement on problems in emotional, cognitive, social functioning, and financial difficulties. On exploring how the caregivers rated differently than the patients, it was found that the former tended to overestimate almost all items, except for problems in social functioning (Table 2). From the group aspect, patients and caregivers had significantly different ratings for physical functioning, role functioning, emotional functioning, and pain (Figure 1).

### Factors Associating with Congruence Level

For patient–physician dyads, we found statistically significant relationships between patient-related variables and congruence in evaluating all three domains. First, age, functional impairment, and burden level together explained 20.3% of the variance in the congruence level of the patient–physician symptom evaluation (*P* = .005). Patients with worse functional impairment, higher burden, and younger age were more likely to be underestimated by the physicians in terms of their symptoms (Figure 2). Second, age, education, symptom severity, and burden level together explained 32.4% of the variance in the congruence level of patient–physician functioning evaluation (*P* = .001). Patients with more severe symptoms, heavier burdens, and younger age were more likely to be underestimated by the physicians in terms of their functioning impairment. Furthermore, physicians were more likely to underestimate functional impairment in patients with low educational levels, compared to those with higher educational levels (Figure 3). Third, functional impairment and HRQOL explained 20.1% of the variance in the congruence level of patient– physician HRQOL evaluation (*P* = .002). Patients’ HRQOL were more likely to be underestimated by the physicians if they had poorer HRQOL and more severe functional impairment. For patient–caregiver dyads, only a significant relationship regarding the HRQOL domain was found; caregivers were likely to underestimate the HRQOL in patients with poorer HRQOL and burden level. The HRQOL and burden level explained 23.1% of the variance in the congruence level of patient–caregiver HRQOL evaluation.

**Figure 2.**
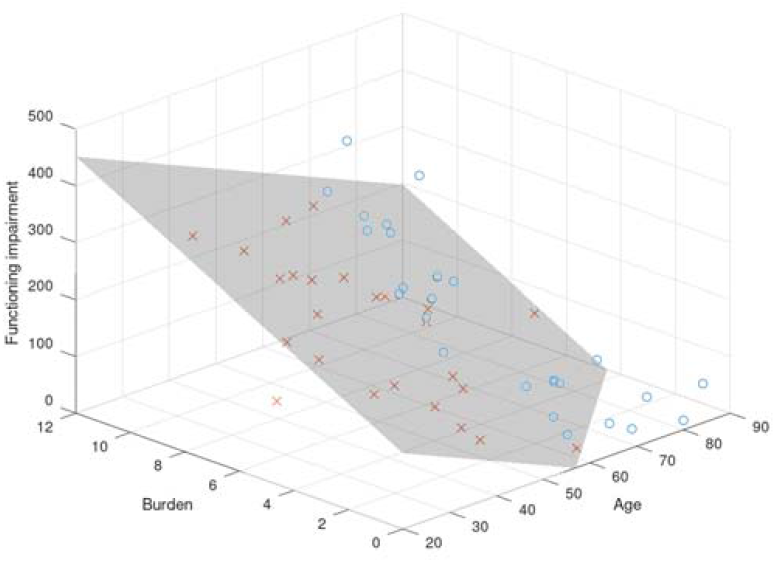
Relationship among Functioning Impairment, Burden Level, Age, and Congruence of Patient-Physician Symptom Evaluation: The gray surface in the 3D regression plot depicts perfect congruence in symptom evaluation (patient-physician difference=0); the circles and crosses demonstrate real data situated above and under the surface, respectively. The space above the surface means overestimation and the space under the surface represents underestimation. Category boundary for functioning impairment is 0-500 and 0-14 for burden.

**Figure 3.**
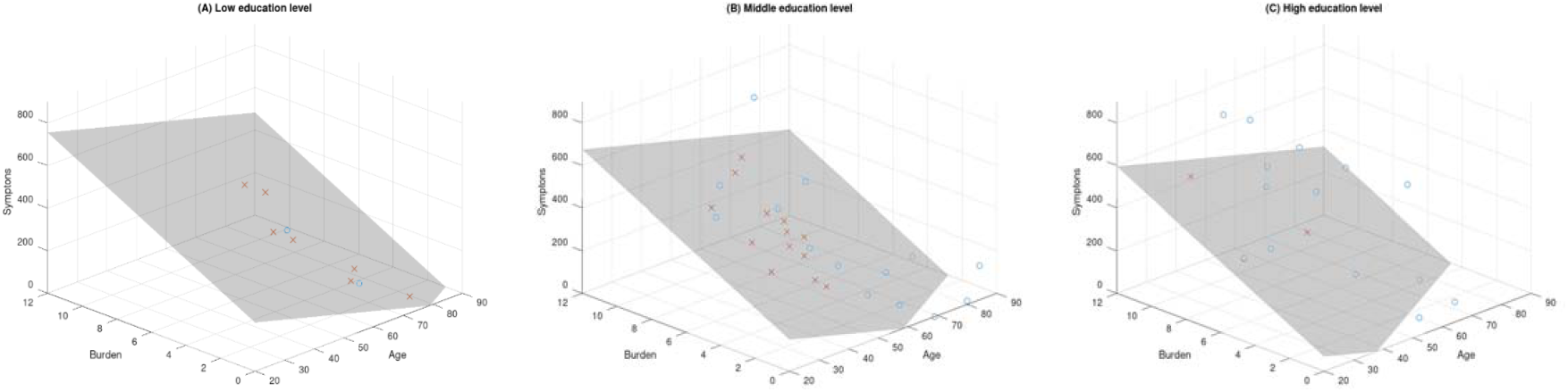
Relationship among Symptom, Burden Level, Age, and Congruence of Patient-Physician Functioning Evaluation according to Three Education Levels: The surfaces in the 3D regression plots depict perfect congruence in functioning evaluation (patient-physician difference=0) according to three education levels: (A) low, (B) middle, and (C) high; the circles and crosses demonstrate real data situated above and under the surface, respectively. The space above the surface means overestimation and the space under the surface represents underestimation. Category boundary for symptoms is 0-900 and 0-14 for burden.

## Data Availability

The authors confirm that the data supporting the findings of this study are available within the article.

## Acknowledgments

We would like to thank palliative shared-care team, department of oncology, and department of hematology-oncology of National Taiwan University Hospital in helping with patient recruitment; and thank Dr. Chin-Hao Chang, National Taiwan University Hospital-Statistical Consulting Unit (NTUH-SCU), and Dr. Yao-Cheng Lin for statistic consultation and figure construction. We appreciate the financial support from Ministry of Science and Technology of Taiwan (grant number: MOST 107-2314-B-002-283-MY2).

